# A scoping review of antimalarial drug resistance markers in Kenya (1987-2022): Toward a National Surveillance Framework and Data Repository

**DOI:** 10.1101/2025.08.16.25333821

**Authors:** Kevin Wamae, John Magudha, Emmanuel Asiimwe, Kariuki Kimani, Regina Kandie, Kibor Keitany, Robert W Snow, L. Isabella Ochola-Oyier

**Affiliations:** KEMRI-Wellcome Trust Research Programme, Kenya; Division of National Malaria Programme, Ministry of Health, Nairobi, Kenya; Centre for Tropical Medicine and Global Health, Nuffield Department of Medicine, University of Oxford, Oxford, UK

**Author notes:** Joint first author (Kevin Wamae, John Magudha).

**Keywords:** Antimalarial drug resistance, surveillance, database

## Abstract

The identification of genetic markers has revolutionised our assessment of antimalarial drug resistance. Tracking the molecular markers of resistance emerged as a valuable tool over 60 years ago, following the identification of sulphadoxine-pyrimethamine genetic resistance markers, *dhfr* and *dhps*. The Preferred Reporting Items for Systematic Reviews and Meta-Analyses extension for Scoping Reviews (PRISMA-ScR) guidelines were used. PubMed/MEDLINE, Embase, Scopus, Google Scholar and Web of Science were systematically searched to identify studies on antimalarial drug resistance markers in Kenya published in English between 01-Jan-1995 and 31-Oct-2024.

The national analysis showed a regional shift in the timelines from the mutant to wild-type *crt* genotype and similarly from the mutant (CVIET) to wild-type (CVMNK) microhaplotype, with the Coast occurring earlier in 2002, while Western Kenya the change occurred later in 2008. MDR1 codons 86 and 1246, also genetic markers of chloroquine resistance have shown a full reversion to the wildtype, that was rising since 1994 in the Coast and 2003 in Western Kenya. By the time drug policy changed to SP in 1999 the dhps mutant genotype was already rising from 1996 in the Coast and 1998 from Western Kenya, while the dhfr codon 108 shift to mutant occurred as early as 1988 in the Coastal parasite populations. Emerging WHO validated k13 mutations were first described, P553L, in 2006 in Kisumu.

This aggregation of data across Kenya demonstrates the utility of this scoping review. The compilation and standardization of over 100 studies provides a high-level, structured overview of when and where resistance markers have been surveyed. This establishes a foundational national repository to support strategic surveillance planning by the Kenya NMCP.

## Introduction

Antimalarial drug resistance (AMDR) poses a major threat to malaria control efforts globally. AMDR refers to the parasites reduced susceptibility to standard antimalarial treatments, leading to prolonged or incomplete parasite clearance that can lead to treatment failure [1]. The malaria parasite has historically evolved to avoid drug action and mutations will continue to emerge to all existing and new antimalarial treatments. The identification of genetic markers of AMDR has revolutionised our assessment of this relentless malaria public health challenge [2–7]. Tracking the molecular markers of resistance emerged as a valuable tool over 60 years ago (with the identification of sulphadoxine-pyrimethamine genetic resistance markers, [3,4]) and allowed an improved understanding of the spread and epidemiology of antimalarial drug resistance.

Chloroquine (CQ) resistance in Africa was at high levels, >90% of the chloroquine resistance transporter (CRT) CVIET mutant haplotype present at the time of widespread clinical failures, and associated with a rise in both malaria morbidity and mortality [8,9]. Following the cessation of CQ use, there was a decline in CVIET parasites across Africa, with some low prevalence (<0.4%) of resistant genotypes reported in some countries, Madagascar in 2007, Malawi in 2009, Zambia in 2013 and Tanzania in 2018 [10]. Conversely, countries that have retained CQ for *P. vivax* treatment (e.g. Ethiopia) continue to have high levels of CVIET mutations [11].

Sulphadoxine-Pyrimethamine (SP), that almost universally replaced CQ as a first-line treatment in the early 2000s across Africa, has shown variable geographic emergence and rates of increase of different mutations to both dihydrofolate reductase (*dhfr*) and dihydropteroate synthase (*dhps*) genotypes [12]. While withdrawn as a treatment, SP continues to be used for the prevention of malaria during pregnancy and infancy and in combination for seasonal malaria chemoprevention. This sustained drug pressure has led to a continued rise in the quintuple ‘super resistant’ *dhfr* and *dhps* combination mutant in East Africa [13], while in West Africa, the quadruple mutant is dominant and the *dhps* 540E mutation is less frequent [14].

Validated WHO artemisinin resistance kelch 13 (*k13*) mutations (WHO) have been detected initially in Southeast Asia [15,16] and since 2015 in East Africa [17]. The initial and widespread, but non-significant mutation A578S (confirmed as not conferring resistance to artemisinin [7]) was described, similarly K189S found outside the propeller domain was also observed in Africa [18]. The current emerging trend of important *k13* mutations in Africa, particularly from the East and Horn of Africa are C469Y/F, R539T, P553L, R561H, R622I and A675V validated artemisinin resistance mutations and P441L and S552C as associated with resistance mutations [19].

National anti-malaria treatment policy dialogues were focused on data from studies of clinical failure during the CQ era. This began to change during the SP era where data on treatment failures was augmented with data from studies of the early identification of *Pfdhfr* and *Pfdhps* mutations [20]. Furthermore, WHO advice on using SP as an intermittent presumptive treatment for prevention is guided by data on mutation rates [21]. International and national antimalarial future drug policy discussions are now driven by molecular surveillance, where the genetic marker of resistance is known, the detection of *Pfk13* mutations, and linked to their clinical association with delayed parasite clearance [22].

The power of genetic markers of AMDR in tracking changes in malaria parasite susceptibility to widely used drugs over time and space has raised the significance of establishing malaria molecular surveillance (MMS) [23]. However, how these enhanced surveillance networks are connected within countries, between countries, data are assembled/shared/standardised and how information are presented in policy forums to guide national decision-making remains under-developed.

MMS in Africa has been reliant on national and international academic research groups with little coordinated national level surveillance or systematic, standardised approaches within countries or regionally. Importantly, data generated at national levels are not always accessible in an easy to understand format to inform action to those charged with making health policy. To address this, we have undertaken a scoping review of AMDR mutations in Kenya, to assemble historical and contemporary data on mutations to previously and currently used antimalarial drugs. The intention being to provide a unique MMS database that can be used by the national malaria control programme (NMCP) to better understand the past, present and possible future of parasite mutations and initiate better future coordination and submission of data assembled by research partners.

## Methods

### Country context

Kenya has a diverse malaria ecology ranging from areas unable to sustain transmission due to altitude/temperature limits on sporogony or semi-arid deserts that cannot support mosquito survival to intense perennial transmission. Traditionally, the most intense transmission has occurred along Kenya’s coastline and around Lake Victoria [24,25]. Despite significant gains in the control of malaria since the launch of Roll Back Malaria nationwide, progress has been less dramatic among lakeside counties in Western Kenya and the southern coastline [24,25]. Since 2010, the Kenyan NMCP has sub-nationally classified the country into five malaria epidemiological zones [26]. These are the lake endemic area-localities around Lake Victoria with stable, high transmission; the coast endemic area-localities along the Indian Ocean coast with low to moderate transmission; the Western highland epidemic prone areas with unstable, year-to-year fluctuations in transmission; the semi-arid seasonal area-northern, eastern, and south-eastern settings with short, acute unpredictable transmission seasons; and the very low-risk area-central highlands and Nairobi, where transmission is either low or absent (Figure 1) [26]. This sub-national stratification has guided the delivery of malaria interventions since 2010, including vector control, early vaccine introduction and the use of intermittent presumptive treatment of malaria in pregnancy with SP: where efforts have been intensified in counties located in the first two endemicity classes around Lake Victoria and along the Indian ocean coast [26]. Early detection, diagnosis and treatment is an important pillar of the malaria strategy, irrespective of endemic classification.

**Figure 1.**
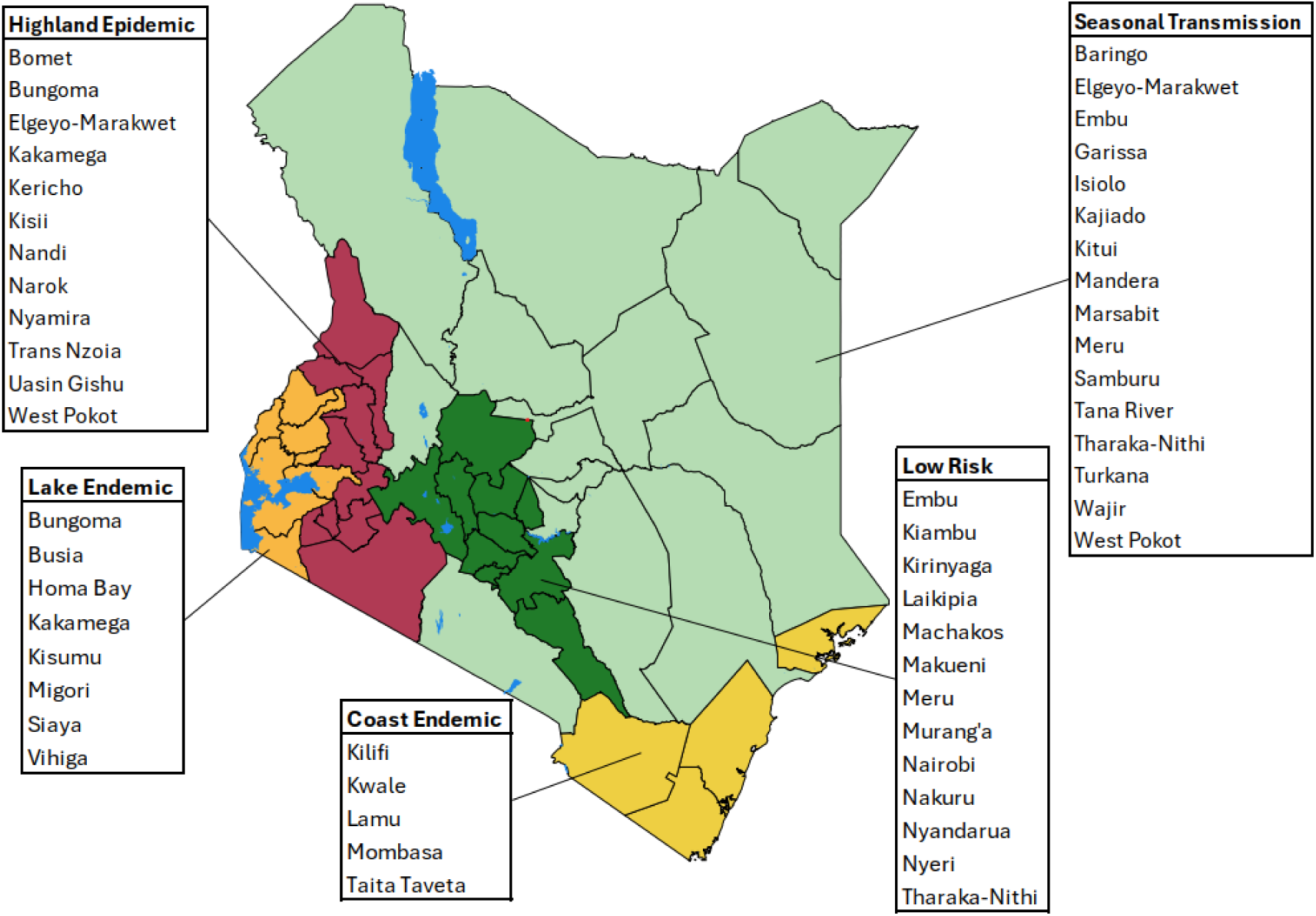
Geographic distribution of malaria epidemiological zones in Kenya. Kenya’s malaria epidemiological zones are defined by the Kenya National Malaria Control Programme. The zones reflect varying transmission patterns: lake endemic (orange) experiences high, stable transmission; coast endemic (yellow) has low to moderate year-round risk; highland epidemic (maroon) has unstable seasonal outbreaks; semi-arid seasonal (green) sees short, acute outbreaks; and low-risk areas (light green) have minimal transmission. Major lakes are in blue, with grouped county labels for each zone (Adapted from [34].

CQ resistance was first reported in Kenya in a tourist in 1978 [27] and in a semi-immune Kenyan in 1982 [28]. Thereafter, escalating treatment failures were documented for over 10 years and the Kenyan Ministry of Health, like many other African countries, were slow to respond [29]. These delays were, in part a result of poor dialogue between local research communities and policy makers [29]. Between 1997-1998, CQ was formally replaced as a first line treatment with SP [30], a decision facilitated by a newly established network of researchers and malaria divisions within the ministries of health in the East Africa region, East African Network for Monitoring Antimalarial Treatment (EANMAT) [31]. This co-created network provided a platform based on mutual trust, ownership of evidence and data sharing across borders that fed directly into policy discussions. EANMAT was subsequently instrumental in providing the evidence to abandon SP in favour of ACTs across the subregion [30,32]. In 2004, Kenya adopted artemether-lumefantrine (AL) as first-line treatment for uncomplicated malaria, which was not implemented effectively until 2006 [30]. Subsequent drug therapeutic and molecular monitoring efforts, mainly supported by the President’s Malaria Initiative (PMI) and the Centres for Disease Control and Prevention (CDC) [33], have been fragmented and reliant on independent research groups, resulting in significant gaps in coordinated national-level surveillance. Hence the need to identify, by means of a scoping review, data that has been collected during and post the EANMAT era.

### Search strategy

The review adheres to the guidelines established by the Preferred Reporting Items for Systematic Reviews and Meta-Analyses extension for Scoping Reviews (PRISMA-ScR) [35].

PubMed/MEDLINE, Embase, Scopus, Google Scholar and Web of Science were systematically searched to identify studies on antimalarial drug resistance markers in Kenya published in English between 01-Jan-1995 and 31-Oct-2024. The search strategy was tailored for each database using a combination of terms and free-text keywords related to Kenya, malaria, drug resistance, and parasite genotyping. Date restrictions and language filters were included where applicable. The complete list of search terms and syntax used for each database is presented in Supplementary table 1.

### Inclusion and Exclusion Criteria

The identified articles were uploaded to Rayyan, a web platform for systematic reviews [36]. Four independent investigators (KW, JM, EA, and KK) reviewed and extracted the data, and disagreements were resolved through discussion and consultation with LIOO. Studies were included if they were conducted in Kenya, were published in English, and focused on molecular markers of resistance in *P. falciparum*. Studies were excluded if they addressed pathogens other than *P. falciparum,* were purely therapeutic efficacy trials or targeted populations beyond the geographical limits of Kenya. Furthermore, studies that did not provide genotype frequencies or consisted solely of reviews of genotyping studies were also excluded. Full-text articles that were not readily accessible were obtained with the librarian’s assistance at the KEMRI-Wellcome Trust Research Programme.

### Screening and Data Extraction

The 110 studies that met the inclusion criteria were systematically reviewed, and data were extracted into comma-separated files (CSV) for each study based on predefined variables (Table 1). A set of R scripts was used to manage and structure the extracted data. Raw allele and microhaplotype frequency data were imported and cleaned. This included parsing fields to extract codon positions, distinguishing wildtype from mutant alleles, and appending standardised gene identifiers from PlasmoDB [37,38]. The cleaned allele and microhaplotype tables were saved for downstream use. Aggregated data from the publications was utilised to generate summary plots of mutation and microhaplotype frequency data by malaria epidemiology zones (Figure 1) and year. Frequencies were either extracted directly or calculated from reported counts, and plots of data availability and temporal trends were generated to guide interpretation. All the extracted variables were compiled into master tables summarising allele and microhaplotype prevalence, resistance profiles and spatial-temporal trends. These data were synthesised to map the distribution of drug resistance markers across Kenya and to identify key gaps in current surveillance.

**Table 1.**
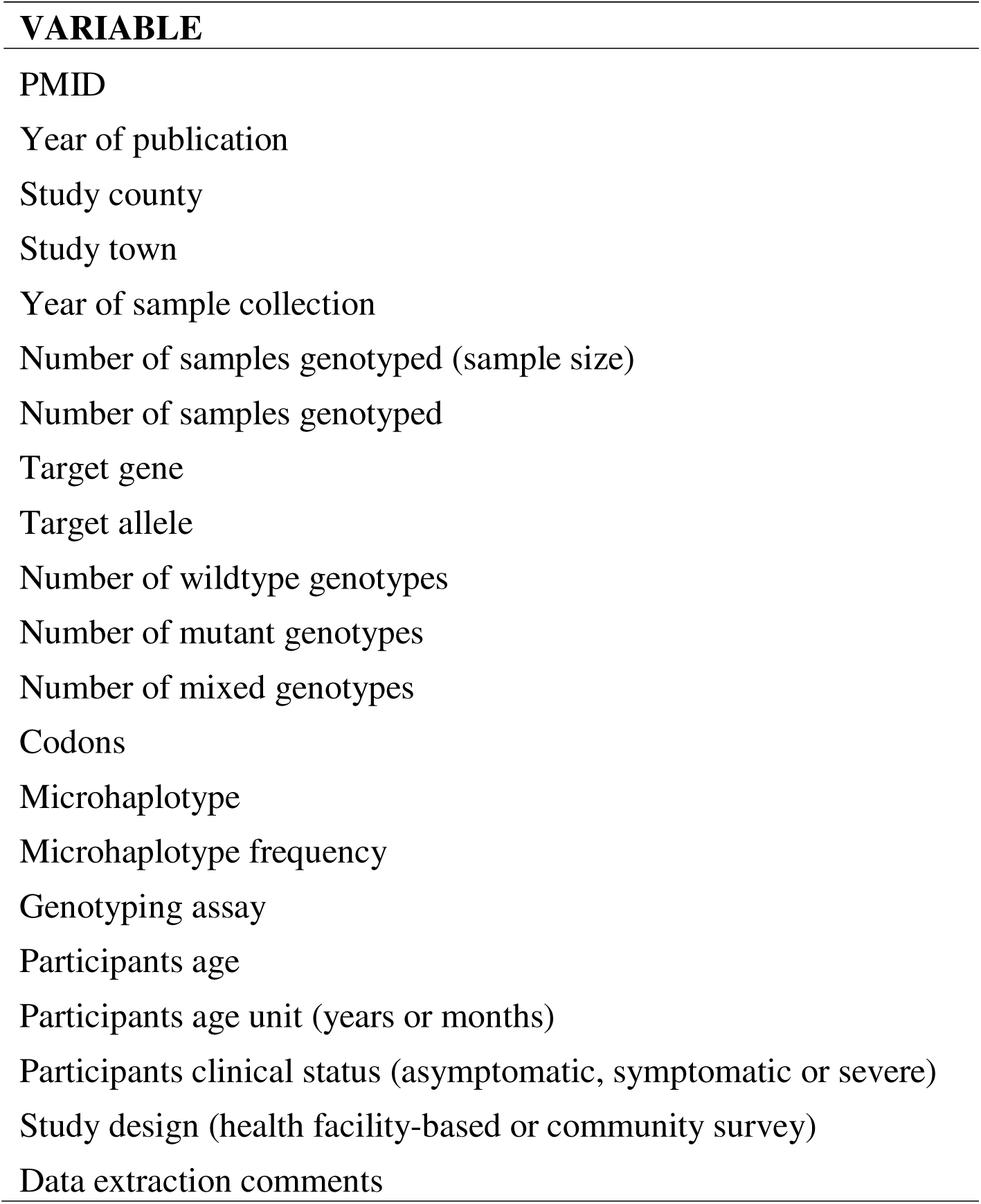
List of variables extracted from the 110 studies.

| **VARIABLE** |
| --- |
| PMID |
| Year of publication |
| Study county |
| Study town |
| Year of sample collection |
| Number of samples genotyped (sample size) |
| Number of samples genotyped |
| Target gene |
| Target allele |
| Number of wildtype genotypes |
| Number of mutant genotypes |
| Number of mixed genotypes |
| Codons |
| Microhaplotype |
| Microhaplotype frequency |
| Genotyping assay |
| Participants age |
| Participants age unit (years or months) |
| Participants clinical status (asymptomatic, symptomatic or severe) |
| Study design (health facility-based or community survey) |
| Data extraction comments |

### Resistance Marker Classification

To support interpretation of the spatiotemporal patterns in antimalarial resistance, we compiled a reference list of key molecular genetic markers (*crt*, *dhfr*, *dhps*, *k13*, *mdr1*) associated with reduced sensitivity to three major drug classes: CQ, SP and ACTs (Table 2). These markers (their associated codons or mutations) form the foundation of molecular surveillance efforts and are critical for assessing the emergence and spread of resistance, guiding treatment policy, and supporting early warning systems. Other putative drug resistance markers were included based on literature providing evidence of their potential role in conferring resistance mainly to ACTs. Other markers such as *ap2-mu, ubp-1*, *exo* and *coronin*, though less well-characterised, are included for completeness, as they have been reported in local studies and may represent early indicators of emerging resistance. This classification framework supports the rationale for gene inclusion in this review and provides a clear reference point for interpreting trends in resistance across time, regions, and drug classes.

**Table 2.** Summarises the most frequently studied resistance molecular markers in malaria-endemic regions.

| Gene | Codon(s) / Mutation(s) | Drug Class | Role in Resistance | Reference |
| --- | --- | --- | --- | --- |
| *crt* | M74I, N75E, K76T, H97Q, A220S, Q271E, N326S, I356T, C350S, R371I | Chloroquine (CQ) | Key mutations conferring CQ resistance | [39–41] |
| *mdr1* | N86Y, Y184F, S1034C, N1042D, D1246Y and copy number variation (CNV) | CQ, Amodiaquine (AQ), Mefloquine (MQ), ACT partner drugs | Modulates CQ and MQ response, lumefantrine tolerance | [42] |
| *dhfr* | N51I, C59R, S108N, I164L | Pyrimethamine (SP) | Core resistance to pyrimethamine | [43–45] |
| *dhps* | S436A, A437G, K540E, A581G, A613S | Sulphadoxine (SP) | Core resistance to sulphadoxine | [46] |
| *k13* | Mutations in the propeller domain | Artemisinin (ACT) | Validated markers of partial resistance | [47] |
| *plasmepsin-II/III* | CNVs | Piperaquine (ACT) | Associated with treatment failure | [48,49] |
| *coronin* | R100K, G50E | ACT | Reduced susceptibility (lab evidence) | [50] |
| *mrp1* | I876V | ACT background | Alters drug efflux | [51] |
| *ap2-mu* | S160N/T | Artemisinin (ACT) | Linked to linked to delayed clearance after ACT treatment | [52] |
| *ubp1* | E1528D, V3275F | ACT background | Modulates artemisinin response | [52,53] |
| *exo* | E415G | Piperaquine (ACT) | Linked to piperaquine failure in Asia | [54] |
| *atp6* | E431K, A623E, S769N | Artemisinin (ACT) | Modulates artemisinin response | [55,56] |
| *falcipain-2a* | S69Stop | Artemisinin (ACT) | Modulates artemisinin response | [57] |
| *nfs* | K65Q | Lumefantrine (ACT) | Lumefantrine tolerance | [58] |
| *arps10* | V127M | Artemisinin (ACT) | Artemisinin background mutations | [59] |
| *mdr2* | T484I | Artemisinin (ACT) | Artemisinin background mutations | [59] |
| *fd* | N193Y | Artemisinin (ACT) | Artemisinin background mutations | [59] |

## Results

### Basic characteristics of included studies

5,986 reports meeting the broad search criteria were identified from the database searches, there were 1,384 duplicate records across the searched databases that were removed. The remaining 4,602 titles and abstracts were screened, resulting in the exclusion of 4,460 records not meeting the scoping review’s inclusion criteria (Figure 2). Of the remaining publications four could not be retrieved. Therefore, the full texts of 138 studies were assessed for eligibility, 29 studies were excluded (2 were from populations outside Kenya and 27 did include genotyping data). 110 studies were included in the review (Figure 2). Studies included were undertaken between 1987 and 2024, with >70% (646/923) being undertaken between 2005 and 2019 (Table 2). Participants across all studies were aged between 1 month to 85 years. 70.8% of reported studies focused on uncomplicated malaria, followed by 13.8% asymptomatic and 1% severe malaria, while 2.1% of cases were not reported. Over two-thirds (62.1%) of the studies were clinic-based, with 11.3% assessing treatment response in therapeutic efficacy studies. Community surveys comprised 20%, while 5.

**Figure 2.**
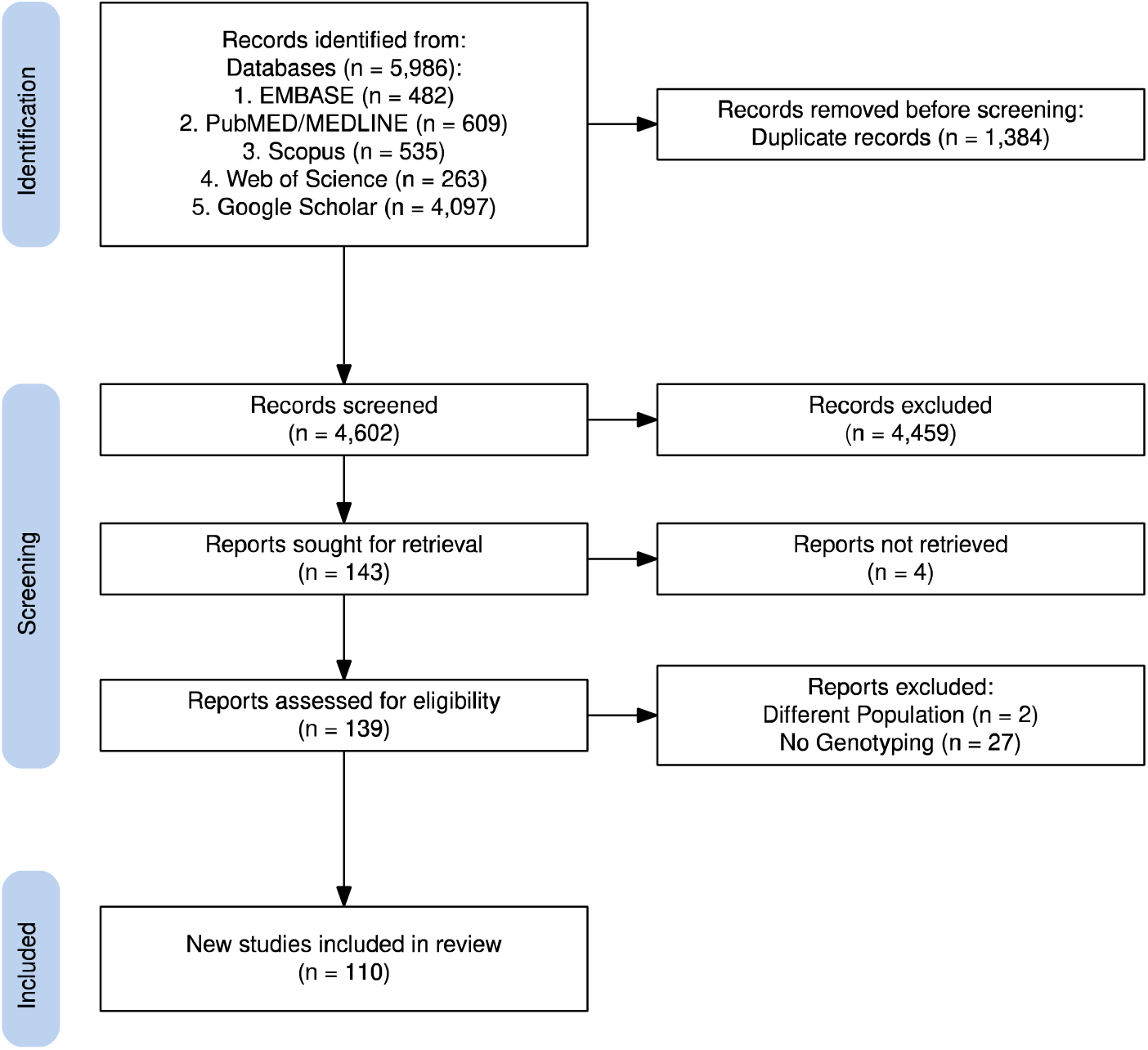
PRISMA Flow Diagram of Study Selection. The figure illustrates the study selection process following PRISMA guidelines. After removing duplicate records, the remaining studies underwent screening based on titles, abstracts, and full texts. Studies that did not meet the eligibility criteria, including those without genotyping data or focusing on non-Plasmodium falciparum pathogens, were excluded. The final number of studies included in the review was 110. This figure was generated using the PRISMA2020 R package v.1.1.3 [60].

Across Kenya’s five malaria epidemiological zones (Figure 1) there were significant variations in data density over the review period. Lakeside and coastal endemic zones characterised by historically high and moderate-to-high malaria transmission had the most comprehensive published molecular surveillance data. Data from the Highland epidemic-prone zones was also present, particularly for the historically important markers, reflecting efforts to monitor areas susceptible to malaria outbreaks (Table 4). Conversely, molecular data were notably sparse and more fragmented in the semi-arid seasonal transmission zones and the low-risk areas (Table 4).

### Molecular surveillance methodologies timeline

The early years (1997-2005) were dominated by Restriction Fragment Length Polymorphism (RFLP) and Sanger sequencing. By 2006, real-time quantitative PCR (qPCR) emerged and maintained consistent usage. From 2012 onwards, next-generation sequencing (NGS) methods, such as amplicon and whole-genome sequencing (WGS), were more prevalent, while SNP genotyping, the MassArray technology, gained traction after 2015, with increased use from 2020 to 2025. More niche applications, Sequence-Specific Oligonucleotide Probe (SSOP) hybridisation assays, SSOP-ELISA, and yeast-expression systems appeared intermittently.

### Nationwide molecular marker surveillance profiles

The data together corroborates the previously published work, with the declining *crt* 76T resistance conferring mutation, and both the Coast and Western Kenya showing a complete reversion to the wildtype, CVMNK genotype, by 2022 (Figure 3A). This is the exact same pattern for resistant mutant amino acids at other genetic loci within the crt gene codons 220, 271 and 371 (Figure 3B). The national analysis shows a regional shift in the timelines for the mutant to wild-type *crt* genotype switches and similarly for the mutant (CVIET) to wild-type (CVMNK) microhaplotype switch (Figure 3C), with the Coast occurring in 2002, while Western Kenya occurred later in 2008.

**Figure 3:**
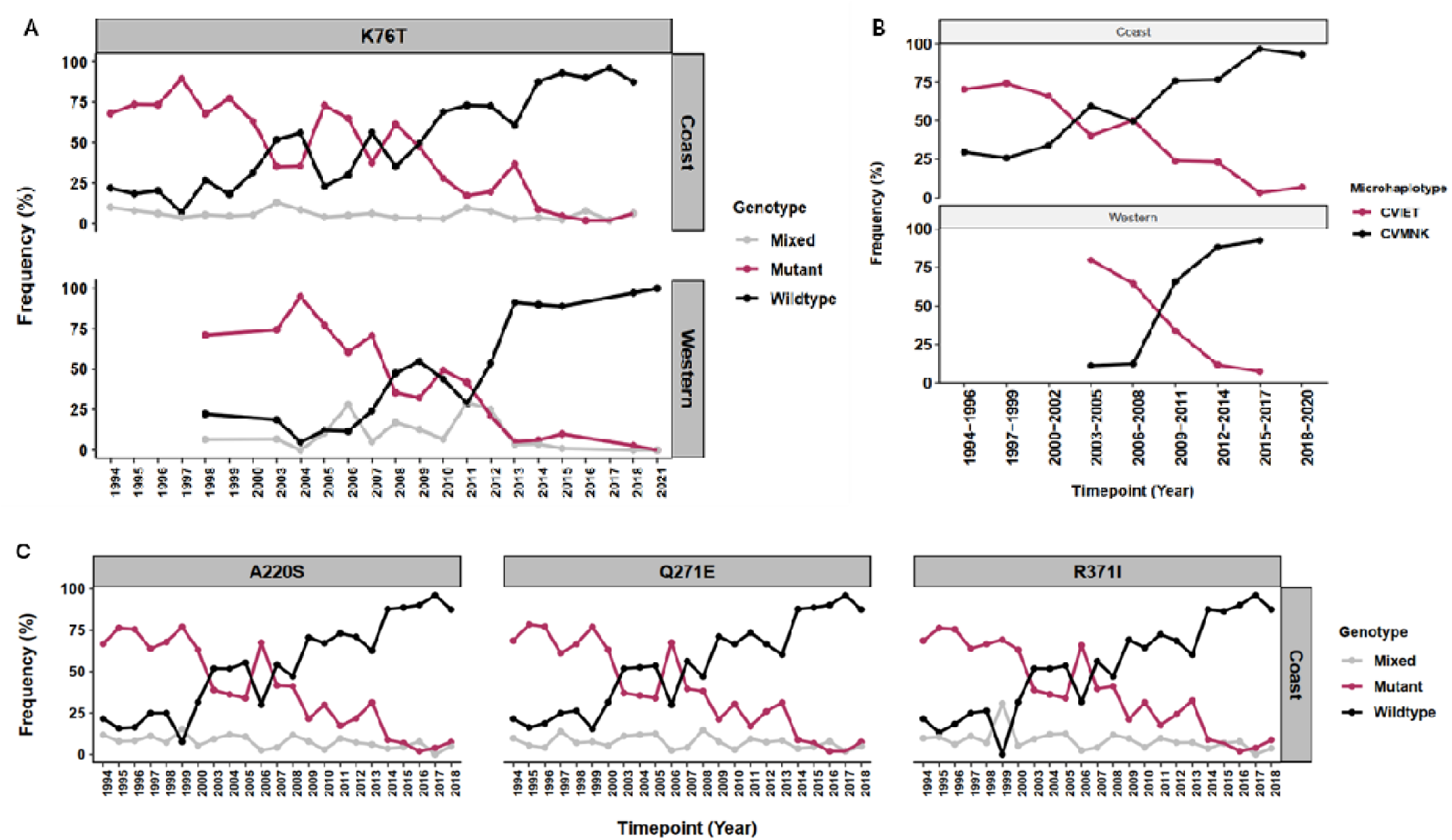
Temporal trends in the frequency of Pfcrt mutations from 1994 to 2018. The (A) K76T; (C) A220S, Q271E, and R371I genotypes; and (B) microhaplotypes CVIET (mutant) and CVMNK (wildtype) are shown. The data was stratified by region: Coast (top row) and Western Kenya (bottom row), for where data was available for both regions. Figure C did not have data from Western Kenya for codons 220, 271 and 371. There was a progressive increase in the frequency of wildtype microhaplotype and alleles were observed at all 4 loci over time.

Furthermore, MDR1 codons 86 and 1246, also genetic markers of chloroquine resistance have shown a full reversion to the wildtype, that was rising since 1994 in the Coast and 2003 in Western Kenya. In contrast, the codon 184 wildtype started dropping in 2006. The NFY and NYD haplotypes together rise in frequency from 2006 and the rare YFY (184 and 1246 double mutant) is only observed in Western Kenya (Figure 4).

**Figure 4.**
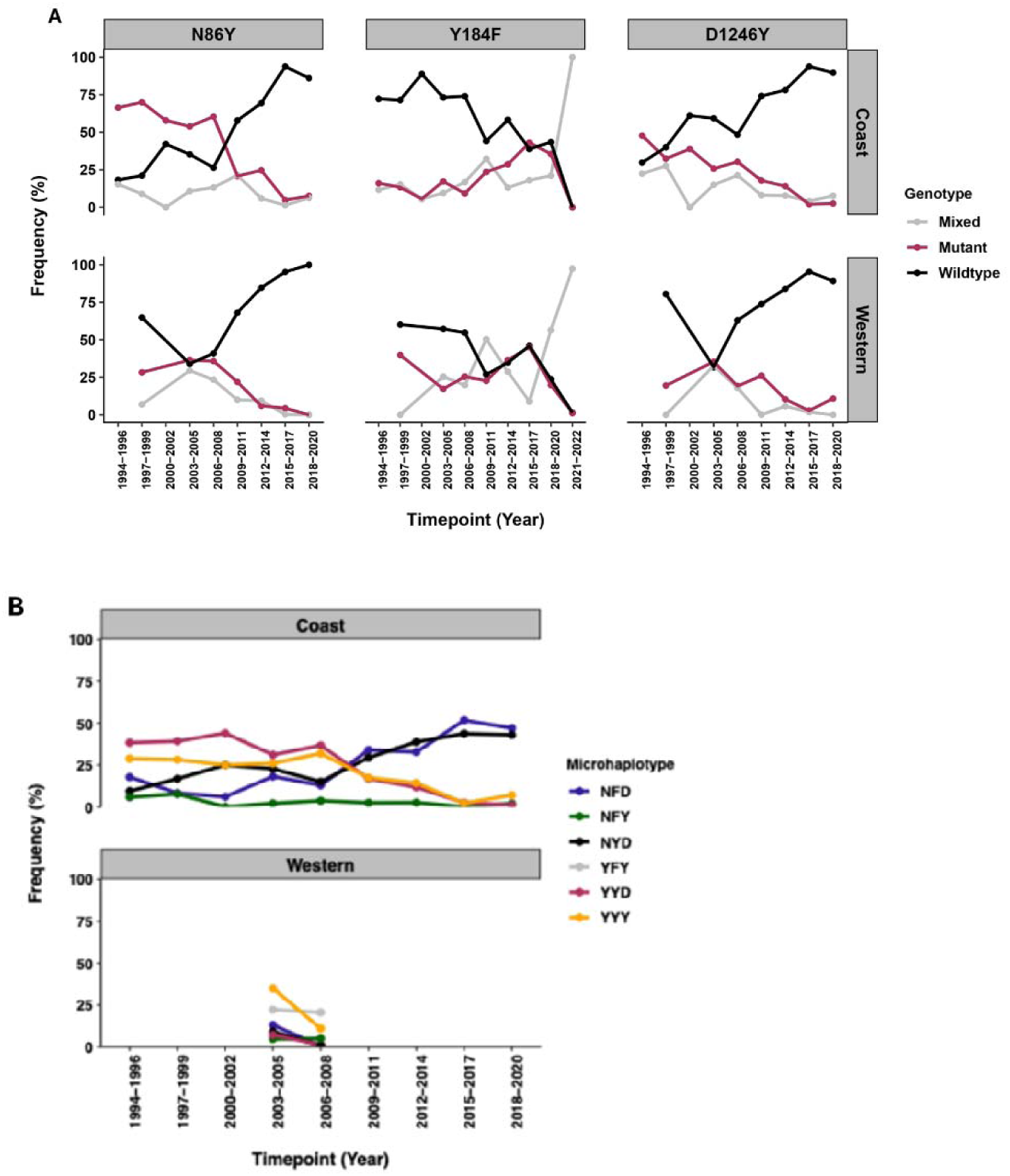
Temporal trends in the frequency of *Pfmdr1* genotypes in Coastal and Western Kenya regions from 1994 to 2022. (A) Each panel shows the proportions of wildtype (black), mutant (red), and mixed (grey) genotypes at three codon positions in Pfmdr1 gene over time. A notable spike in mixed Y184F genotypes during 2021–2022 in Western Kenya (100%) resulted from the fact that only one study [61] contributed data during this timepoint, and majority of the samples from that dataset were mixed infections. No data was available for codons N86Y and D1246Y beyond 2020. (B) The NFD microhaplotype, associated with reduced lumefantrine susceptibility, has shown a steady increase in Coastal Kenya at similar proportions to the wildtype, NYD microhaplotype. The triple mutant YFY was only detected at low frequencies in Western Kenya between 2003 and 2008. Other haplotypes, including NFY, YYD, and YYY, remained at low prevalence over time.

For *dhps*, by the time drug policy changed to SP in 1999 the mutant was already rising from 1996 in the Coast and 1998 from Western Kenya (based on available data) (Figure 5A). The mutant genotype was already taking over as the dominant genotype in both regions of th country, clearly demonstrated in the dhps haplotype analysis from 1994 with a wild type to mutant shift by 1999 (Figure 5B). Of note, across both regions of the country codons 436, 581 and 613 were predominantly wildtype over time.

**Figure 5.**
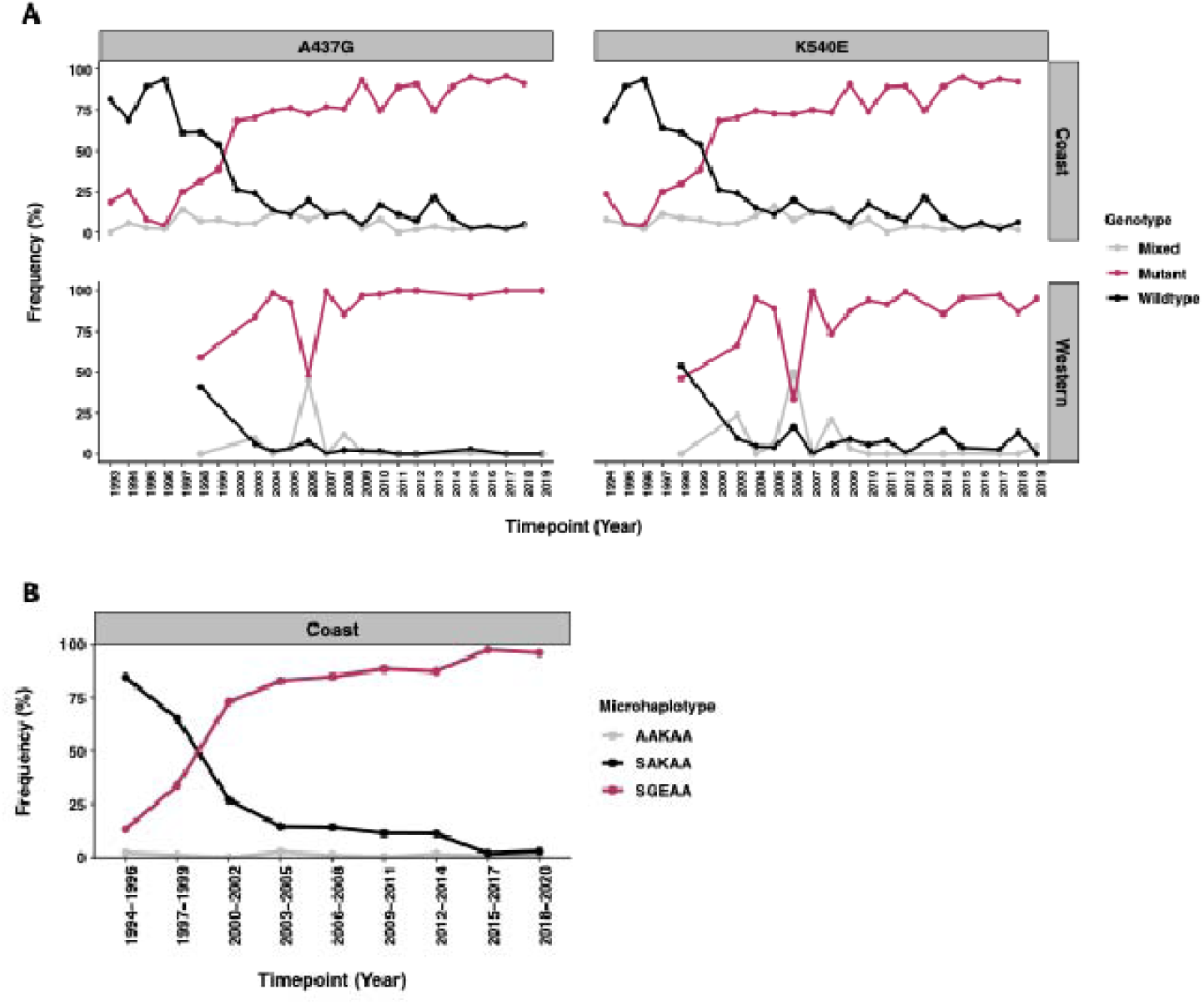
The temporal trends in the dhps alleles, marker for sulphadoxine resistance. The mutant 437G and 540E genotypes remained near fixation from 2016. However, a notable dip in their frequencies was observed in 2006, primarily due to a high proportion of mixed genotypes reported at that timepoint. The dhps SGEA double mutant microhaplotype, characterized by the alleles at codons S436, 437G, 540E, and A581, steadily increased over time in the coastal region, reaching near fixation by 2015. In contrast, the wild-type SAKA haplotype declined sharply over time to 0% also by 2015. A remarkable shift between these haplotypes took place between 1999 and 2000, coinciding with the replacement of chloroquine with sulphadoxine-pyrimethamine.

For the pyrimethamine resistance marker, *dhfr*, as early as 1988, codon 108 started to shift to the mutant genotype in the Coastal parasite populations, codons 51 and 59 shifted later in 1996 (Figure 6A), also reflected in the triple mutant IRNI haplotype in the Coast (Figure 6B). The Western Kenya data was patchy and thus an assessment of the earliest time points the mutations rose in frequency could not be made and there was no microhaplotype data apart from 2006-2008. Furthermore, there has a high reporting of mixed infections in 2008 compared to all other time points, resulting in a drastic drop in the mutant frequencies in Western Kenya. From 2018, the wildtype amino acid at codon 164 showed a decline in frequency in Western Kenya only, where genotyping data is available post-2018 (Figure 6A).

**Figure 6.**
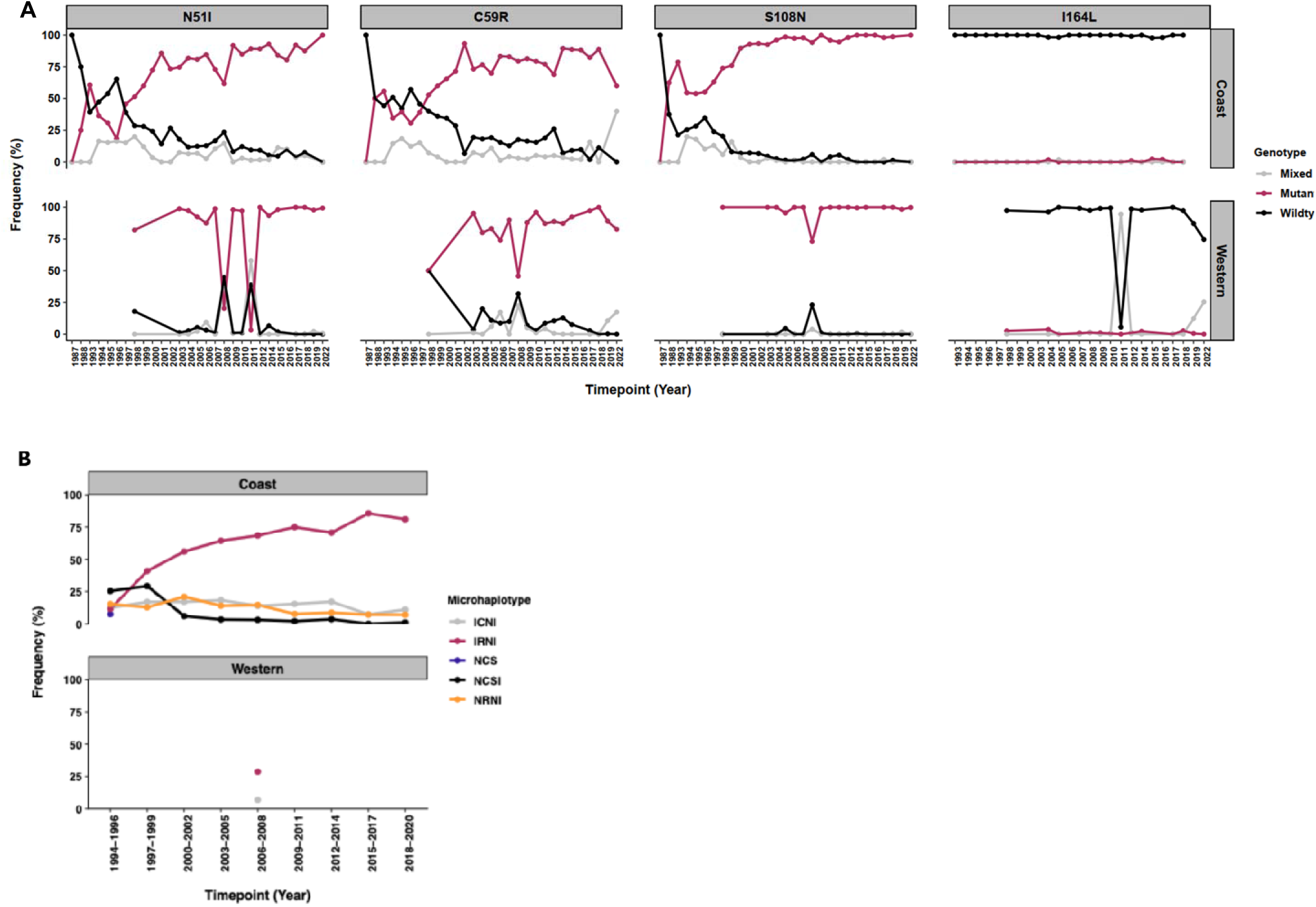
Temporal trends in dhfr alleles across Coastal and Western Kenya regions. (A) The switch from wildtype to mutant alleles occurred as early as 1998 for 3 loci, 51, 59 and 108. Codons 51I and 108N mutations were near fixation in both regions of the country. Due to incomplete data, the 2013 timepoint for Western Kenya was excluded from the plot but is available in Supplementary Table 1. furthermore, in Western Kenya, some studies reported a high number of mixed infections that altered the temporal trend, highlighting the need for consistency in defining mutations across studies and time. The codon 164L mutation is beginning to emerge in mixed infections in Western Kenya revealing the reduction in wildtype parasites, warranting continued surveillance. (B) The temporal trends in dhfr microhaplotypes. In Coastal Kenya, a distinct shift from the wildtype NCSI to the triple mutant IRNI microhaplotype occurred from 1996. The IRNI has steadily increased in frequency, approaching fixation in the later time periods. The wildtype is rare and mostly absent since 2003 along the Coast. In Western Kenya, temporal trends could not be assessed due to missing data for several key years.

A common K13 mutation across time was the A578S but at a low frequency (<5%). Emerging WHO validated k13 mutations were first described, P553L, in 2006 in Kisumu at 2% (Table 3). However, since 2022 the observations of the validated mutations, C469Y, P553L and A675V, was geographically widespread across Western Kenya. There were no observations of these mutations along the Coast, apart from a frequent observation of a synonymous change at codon C469C from 1998 in Kilifi and observations across counties in Western Kenya to 2022 and a single observation of R539K in 2013 in Malindi (Supplementary Table 2). There was a WHO mutation associated with resistance, V568G in 2003 and 2013 in Kisumu at low frequency 5.4% and 2.4%, respectively. Similarly, S552C was observed in 1994 in Kilifi, another WHO mutation associated with resistance (Supplementary Table 2).

**Table 3:** Demographic Characteristics of Study Participants.

|  | Central<br>(N=2) | Coast<br>(N=78) | Eastern<br>(N=1) | Rift Valley<br>(N=11) | Western<br>(N=88) | Multiple<br>(N=14) | Unknown<br>(N=1) | Overall<br>(N=195) |
| --- | --- | --- | --- | --- | --- | --- | --- | --- |
| **Age Group** |  |  |  |  |  |  |  |  |
| ≤5 | 1 (50.0%) | 17 (21.8%) | 0 | 4 (36.4%) | 34 (38.6%) | 1 (7.1%) | 1 (100%) | 58 (29.7%) |
| ≤15 | 0 | 23 (29.5%) | 1 (100%) | 0 (0%) | 12 (13.6%) | 0 | 0 | 36 (18.5%) |
| >15 | 1 (50.0%) | 9 (11.5%) | 0 | 5 (45.5%) | 14 (15.9%) | 12 (85.7%) | 0 | 41 (21.0%) |
| Missing | 0 | 29 (37.2%) | 0 | 2 (18.2%) | 28 (31.8%) | 1 (7.1%) | 0 | 60 (30.8%) |
| **Participants Clinical Case** |  |  |  |  |  |  |  |  |
| Uncomplicated | 2 (100%) | 52 (66.7%) | 0 | 9 (81.8%) | 61 (69.3%) | 13 (92.9%) | 1 (100%) | 138 (70.8%) |
| Asymptomatic | 0 | 3 (3.8%) | 0 | 1 (9.1%) | 22 (25.0%) | 1 (7.1%) | 0 | 27 (13.8%) |
| Asymptomatic,uncomplicated* | 0 | 23 (29.5%) | 0 | 0 (0%) | 1 (1.1%) | 0 | 0 | 24 (12.3%) |
| Severe | 0 | 0 | 1 (100%) | 0 (0%) | 1 (1.1%) | 0 | 0 | 2 (1.0%) |
| Missing | 0 | 0 | 0 | 1 (9.1%) | 3 (3.4%) | 0 | 0 | 4 (2.1%) |
| **Study Design** |  |  |  |  |  |  |  |  |
| Clinic | 2 (100%) | 50 (64.1%) | 1 (100%) | 8 (72.7%) | 52 (59.1%) | 7 (50.0%) | 1 (100%) | 121 (62.1%) |
| Clinic/community survey | 0 | 23 (29.5%) | 0 | 0 | 0 | 0 | 0 | 23 (11.8%) |
| Community survey | 0 | 4 (5.1%) | 0 | 2 (18.2%) | 29 (33.0%) | 4 (28.6%) | 0 | 39 (20.0%) |
| Vaccine trial (clinic)/community survey* | 0 | 1 (1.3%) | 0 | 0 | 0 | 0 | 0 | 1 (0.5%) |
| Missing | 0 | 0 | 0 | 1 (9.1%) | 7 (8.0%) | 3 (21.4%) | 0 | 11 (5.6%) |

‘\*’ indicates studies that collected samples across multiple classification categories, either multiple clinical case definitions or multiple study design types; ‘multiple’ refers to the studies that include data from an aggregate of counties across different regions of the country; ‘missing’ indicates data not available in the study and ‘unknown’ the sdenotes that the tudy did not provide detail on county or region.

**Table 4.** Number of studies sampling across time periods and malaria zones in Kenya.

| Marker | Malaria Zone | Sampling Timepoints |  |  |  |  |  |  |
| --- | --- | --- | --- | --- | --- | --- | --- | --- |
|  |  | 1987-1993 | 1994-1999 | 2000-2004 | 2005-2009 | 2010-2014 | 2015-2019 | 2020-2024 |
| *crt* | Lake endemic | 0 | 6 | 5 | 20 | 20 | 8 | 0 |
|  | Highland epidemic prone | 0 | 2 | 1 | 17 | 6 | 4 | 0 |
|  | Coast endemic | 1 | 10 | 3 | 14 | 7 | 7 | 0 |
|  | Semi-arid seasonal transmission | 0 | 1 | 0 | 3 | 0 | 1 | 0 |
| *mdr1* | Lake endemic | 0 | 8 | 6 | 22 | 22 | 12 | 6 |
|  | Highland epidemic prone | 0 | 2 | 1 | 19 | 10 | 6 | 3 |
|  | Coast endemic | 1 | 9 | 3 | 15 | 13 | 8 | 1 |
|  | Low-risk | 0 | 0 | 0 | 0 | 0 | 1 | 0 |
|  | Semi-arid seasonal transmission | 0 | 2 | 0 | 3 | 1 | 1 | 1 |
| *dhfr* | Coast endemic | 9 | 16 | 9 | 13 | 10 | 5 | 1 |
|  | Lake endemic | 1 | 4 | 9 | 12 | 15 | 13 | 6 |
|  | Highland epidemic prone | 0 | 3 | 1 | 13 | 6 | 6 | 4 |
|  | Semi-arid seasonal transmission | 0 | 2 | 0 | 1 | 0 | 1 | 1 |
| *dhps* | Lake endemic | 1 | 4 | 6 | 12 | 14 | 13 | 6 |
|  | Highland epidemic prone | 0 | 2 | 1 | 13 | 6 | 6 | 4 |
|  | Coast endemic | 3 | 11 | 3 | 9 | 10 | 6 | 1 |
|  | Semi-arid seasonal transmission | 0 | 2 | 0 | 1 | 0 | 1 | 1 |
| *k13* | Lake endemic | 0 | 2 | 1 | 5 | 8 | 11 | 7 |
|  | Coast endemic | 0 | 7 | 2 | 6 | 7 | 6 | 1 |
|  | Highland epidemic prone | 0 | 1 | 1 | 1 | 2 | 6 | 5 |
|  | Semi-arid seasonal transmission | 0 | 0 | 0 | 0 | 1 | 0 | 1 |
|  | Low-risk | 0 | 0 | 0 | 0 | 0 | 1 | 0 |
| *ap2-mu* | Lake endemic | 0 | 0 | 0 | 1 | 3 | 1 | 0 |
|  | Coast endemic | 0 | 2 | 0 | 1 | 1 | 1 | 0 |
| *arps10* | Coast endemic | 0 | 6 | 3 | 5 | 5 | 4 | 0 |
| *atp6* | Lake endemic | 0 | 1 | 0 | 0 | 0 | 0 | 0 |
| *coronin* | Lake endemic | 0 | 0 | 0 | 0 | 0 | 2 | 0 |
| *exo* | Coast endemic | 0 | 6 | 3 | 5 | 5 | 4 | 0 |
| *falcipain-2a* | Coast endemic | 0 | 2 | 0 | 1 | 1 | 1 | 0 |
| *fd* | Coast endemic | 0 | 6 | 3 | 5 | 5 | 4 | 0 |
| *mdr2* | Coast endemic | 0 | 8 | 3 | 6 | 6 | 5 | 0 |
| *mrp1* | Lake endemic | 0 | 0 | 1 | 3 | 3 | 2 | 0 |
|  | Highland epidemic prone | 0 | 0 | 0 | 4 | 4 | 2 | 0 |
|  | Semi-arid seasonal transmission | 0 | 0 | 0 | 2 | 2 | 1 | 0 |
|  | Coast endemic | 0 | 0 | 0 | 2 | 2 | 1 | 0 |
| *nfs1* | Lake endemic | 0 | 0 | 0 | 0 | 0 | 3 | 0 |
|  | Coast endemic | 0 | 2 | 0 | 1 | 1 | 2 | 0 |
| plasmepsin-II | Lake endemic | 0 | 0 | 0 | 0 | 1 | 5 | 0 |
|  | Coast endemic | 0 | 6 | 3 | 5 | 6 | 6 | 0 |
|  | Highland epidemic prone | 0 | 0 | 0 | 0 | 2 | 2 | 0 |
|  | Semi-arid seasonal transmission | 0 | 0 | 0 | 0 | 1 | 1 | 0 |
| plasmepsin-III | Coast endemic | 0 | 6 | 3 | 5 | 6 | 6 | 0 |
|  | Lake endemic | 0 | 0 | 0 | 0 | 1 | 1 | 0 |
|  | Highland epidemic prone | 0 | 0 | 0 | 0 | 2 | 2 | 0 |
|  | Semi-arid seasonal transmission | 0 | 0 | 0 | 0 | 1 | 1 | 0 |
| ubp-1 | Lake endemic | 0 | 0 | 0 | 1 | 1 | 0 | 0 |
|  | Coast endemic | 0 | 2 | 0 | 1 | 1 | 1 | 0 |
| Total number of studies per year |  | 16 | 141 | 71 | 247 | 218 | 181 | 49 |

**Table 5.** List of WHO validated K13 mutations in Kenya.

| Sampling year | County | Malaria epidemiological zones | Sample size | N458Y | C469Y | R539T | P553L | R561H | A675V | Reference |
| --- | --- | --- | --- | --- | --- | --- | --- | --- | --- | --- |
| 2006 | Kisumu | Lake endemic | 50 | - | - | - | 2 | - | - | [62] |
| 2018-2022 | Kisumu, Kisii, Kakamega, Homa Bay | Lake & highland epidemic | 775 | - | - | - | - | - | 0.4 | [63] |
| 2021 | Kisii | Highland epidemic | 13 | 20 | - | 20 | - | 40 | - | [64] |
| 2022 | Bungoma | Highland epidemic | 74 | - | 5.9* | - | - | - | - | [61] |
| 2022 | Busia | Lake endemic | 322 | - | 2.8* | - | - | - | - | [61] |
| 2022 | Busia | Lake endemic | 226 | - | 4.8 | - | - | - | - | [65] |
| 2022 | Kakamega | Highland epidemic | 145 | - | - | - | 4.3* | - | - | [61] |
| 2022 | Kisumu | Lake endemic | 161 | - | 10.7* | - | 3.6* | - | 7.1* | [61] |
| 2022 | Migori | Lake endemic | 275 | - | 2.1* | - | - | - | - | [61] |
| 2022 | Siaya | Lake endemic | 337 | - | 6.9* | - | - | - | - | [61] |
| 2022 | Turkana | Seasonal transmission | 92 | - | 4.3* | - | - | - | - | [61] |
| 2022 | Vihiga | Lake endemic | 68 | - | 2.1* | - | - | - | - | [61] |
| 2022 | West Pokot | Seasonal transmission | 32 | - | 33.3* | - | 14.3* | - | 50* | [61] |

An asterisk (\*) denotes mixed genotype frequencies, where both wild-type and mutant alleles were detected within the same sample. Dash (-) indicates that the mutation was not detected or not assessed in the study.

There was data on other putative drug resistance markers (based on previous publications, Table 1, Supplementary table 3) that showed no change in allele frequency trends over time (ap2mu, atp6, falcipain-2a, mrp1, nfs and ubp1) and those that were 100% wildtype (arps10, exo, fd and mdr2). However, for coronin, only one study reported data over a two-year period; therefore, no trend analysis was performed. The data is available in Supplementary Table 1.

## Discussion

This scoping review identified data variables that can be utilized to generate a shared data standard for the country. It can be populated as an aggregate dataset that provides a nationwide overview of the presence and frequency of mutations over time. Of immediate importance is the continued presence of WHO validated k13 mutations in Western Kenya, from Turkana southwards to Homa Bay and Migori counties [61,63,65]. Due to the low numbers of samples (and reported high frequency) from Maniga and colleagues[64], the sequence data available on GenBank was reassembled to countercheck the reported frequencies, our alignment analysis was not able to verify the presence of the reported SNPs. However, this review highlights the early identification of a now WHO validated mutation, P553L, in a study published in 2006 [62]. Additionally, of an emerging synonymous change, C469C, in 1998 [66] at a now WHO validated locus resulting in a C469Y change. Both are important additions towards understanding the timelines of genetic changes at the k13 locus nationally.

An additional limitation was the reporting of mixed and mutant infections across studies; this requires standardization to support robust temporal and spatial comparisons. Mixed infections may sometimes be referred to as mutants since they are likely to lead to a resistance phenotype. However, there was also the observed change in molecular methodology, from the traditional to advanced and newer technologies. These each require a clear definition for calling mixed infections, from the presence of multiple bands by RFLP, to a peak within a peak by capillary sequencing and a read cut-off to determine the limit of detection of low frequency variants by NGS. The use of NGS was partly driven by the need for high-throughput genotyping and with it comes a concerted effort to standardize methodologies, data and analytical pipelines for an easy-to-understand output for the policy makers.

Other important potentially emerging mutations that require continued monitoring are the *dhfr* 164L mutation that will make the parasites super-resistant to SP and for which data is required from the Coast; and the *dhps* 581G mutation, known to be associated or to arise due to intermittent preventive treatment in pregnancy (IPTp) [67], that is still wildtype. The *dhfr* and *dhps* genes should continue to be tracked in Western Kenya, given the limited data in this review, where the malaria burden is highest and SP in IPTp is a major malaria control measure. The early shift to the mutant genotypes for *dhps* and *dhfr* reflects directional selection pressure exerted by widespread SP use, driving the expansion of the resistant parasite populations.

CQ has reverted to a >99.9% wild type population across Kenya. The delays in resistance switches between the East and West of the country highlight the potential lack of uniformity in the complete withdrawal of chloroquine in the late 90s during the national policy change to SP. These are lessons that are useful for any future drug regimen changes and an important indicator to work closely with the private sector on national treatment changes.The *crt* gene and variant (76T) is the primary mediator of CQR, increasing the export of CQ from the food vacuole away from its target haem [68]. Additionally, MDR1 modulates the parasites to several antimalarial drugs, including CQ, it is involved in the hydrophobic antimalarial efflux [69]. Additional mutations following the crt temporal pattern, codons 220, 271 and 371, were described in the Coastal dataset and is missing from Western Kenya, a gap that requires data. These loci were identified by whole genome sequencing, showing the value of NGS to identify additional loci. The current complement of 7 crt point mutations along the Coast are from codons 72, 74, 75, 76, 220, 271 and 371, which is consistent with the pattern for parasites from Africa that also includes codon 236 [39] that was wildtype. Furthermore, the South American combination includes codon 356 [39], which was also wild-type in the current dataset. The two Pfmdr1 mutations 86Y and 1246Y (found in Africa) mediate the parasite’s decreased susceptibility to chloroquine and amodiaquine, but increased sensitivity to lumefantrine, mefloquine, and artemisinins [70–72]. These mutations followed the K76T pattern of a reduction of the mutation alleles over time. Additional mutations 1034C and 1042D (observed outside Africa) have been associated with altered sensitivity to lumefantrine, mefloquine, and artemisinins [71,73–75]. These mutations were not observed in Kenya as these loci were primarily wildtype.

The majority of long-term data was generated from the Coast and Western Kenya. This is not surprising, since these two regions are locations for national research partners working on malaria with long-term investment in research infrastructure and international collaborations (e.g., KEMRI collaborative centres in Kisumu and Kilifi).The semi-arid seasonal transmission zones and the low-risk areas have traditionally been neglected by the malaria research communities in Kenya, due to the low infection rates and disease burden and despite these regions having frequent malaria outbreaks. These geographical gaps in data are important to redress through future surveillance efforts as the epidemiology of mutation rates could be different in low transmission, semi-arid, pastoralist areas bordering other countries for a more comprehensive national data repository.

This aggregation of data across Kenya demonstrates the utility of this scoping review. In isolation, based on the focus of the research institutions working in Western Kenya and along the Coast, the molecular data corroborated the national changes in antimalarial drug policy. Furthermore, it demonstrates the power of a consolidated database and the essential variables to support data sharing (Table 1). The compilation and standardization of over 100 studies provides a high-level, structured overview of when and where resistance markers have been surveyed. This establishes a foundational national repository to support strategic surveillance planning by the Kenya NMCP. It further highlights the urgent need for more equitable surveillance efforts to ensure national representation. A centralised MMS repository will allow for the necessary resources and technical support to be mobilised; expertise to be shared by developing a network of laboratories for genomics and bioinformatics; standardised protocols to generate reproducible data and enable reagents sourcing at scale; and importantly coordinated sample referral systems [76]. For malaria this is important as control interventions can be targeted to regions where resistance is emerging as evidenced from the current data in Western Kenya that also highlights the need for border control interventions. This has been shared with the NMCP as a policy brief/ technical report for researchers in the country to populate in near real-time to support the national MMS efforts. It will further aid the tracking of drug resistance mutations while identifying potential hotspot areas of the country where resistance mutations are emerging.

## Declarations

### Ethics approval and consent to participate

Not applicable.

### Consent for publication

All authors read the final version and agree to publication.

### Availability of data and materials

All data generated or analysed during this study are included in this published article and its supplementary information files.

### Authors’ contributions

LIOO and RWS conceived and designed the study. KW and JM conducted the literature search. KW, JM, EA, and KK were responsible for data acquisition, screening of primary articles, and data extraction. KW and JM performed the data analysis and generated the summary outputs. KW, JM, EA, and KK drafted the initial manuscript. All authors (KW, JM, EA, KK, RK, KK, RWS, and LIOO) contributed to the interpretation of findings, provided critical revisions, and approved the final version of the manuscript

## Acknowledgements

We acknowledge the contributions of Alex Maina for his support in downloading the research articles. We also thank all authors of the original studies included in this review, whose work forms the foundation of this synthesis

## References

1. WHO. Global Report on Antimalarial Drug Efficacy and Drug Resistance: 2000-2010. World Health Organisation. http://www.who.int/malaria/publications/atoz/9789241500470/en/; 2010.

2. Wootton JC, Feng X, Ferdig MT, Cooper RA, Mu J, Baruch DI, et al. Genetic diversity and chloroquine selective sweeps in Plasmodium falciparum. Nature [Internet]. 2002 Jul;418(6895):320–3. Available from: http://www.nature.com/articles/nature00813

3. Roper C, Pearce R, Nair S, Sharp B, Nosten F, Anderson T. Intercontinental spread of pyrimethamine-resistant malaria. Science (1979) [Internet]. 2004 Aug 20;305(5687):1124. Available from: http://www.sciencemag.org/cgi/doi/10.1126/science.1098876

4. Mita T, Venkatesan M, Ohashi J, Culleton R, Takahashi N, Tsukahara T, et al. Limited geographical origin and global spread of sulfadoxine-resistant dhps alleles in plasmodium falciparum populations. Journal of Infectious Diseases. 2011;204(12):1980–8.

5. Ashley EA, Dhorda M, Fairhurst RM, Amaratunga C, Lim P, Suon S, et al. Spread of Artemisinin Resistance in *Plasmodium falciparum* Malaria. New England Journal of Medicine [Internet]. 2014 Jul 31;371(5):411–23. Available from: http://www.nejm.org/doi/abs/10.1056/NEJMoa1314981

6. MalariaGEN Plasmodium falciparum Community Project. Genomic epidemiology of artemisinin resistant malaria. Elife [Internet]. 2016 Mar 4;5(MARCH2016):1–29. Available from: http://elifesciences.org/lookup/doi/10.7554/eLife.08714

7. Ménard D, Khim N, Beghain J, Adegnika AA, Shafiul-Alam M, Amodu O, et al. A Worldwide Map of Plasmodium falciparum K13-Propeller Polymorphisms. New England Journal of Medicine [Internet]. 2016 Jun 23;374(25):2453–64. Available from: http://www.nejm.org/doi/10.1056/NEJMoa1513137

8. Trape JF, Pison G, Preziosi MP, Enel C, du Loû AD, Delaunay V, et al. Impact of chloroquine resistance on malaria mortality. Comptes Rendus de l’Académie des Sciences - Series III - Sciences de la Vie [Internet]. 1998 Aug;321(8):689–97. Available from: https://linkinghub.elsevier.com/retrieve/pii/S0764446998800097

9. Korenromp EL, Williams BG, Gouws E, Dye C, Snow RW. Measurement of trends in childhood malaria mortality in Africa: An assessment of progress toward targets based on verbal autopsy. Lancet Infectious Diseases. 2003;3(6):349–58.

10. Njiro BJ, Mutagonda RF, Chamani AT, Mwakyandile T, Sabas D, Bwire GM. Molecular surveillance of chloroquine-resistant *Plasmodium falciparum* in sub-Saharan African countries after withdrawal of chloroquine for treatment of uncomplicated malaria: A systematic review. J Infect Public Health [Internet]. 2022 May;15(5):550–7. Available from: https://linkinghub.elsevier.com/retrieve/pii/S1876034122000715

11. Fola AA, Feleke SM, Mohammed H, Brhane BG, Hennelly CM, Assefa A, et al. *Plasmodium falciparum* resistant to artemisinin and diagnostics have emerged in Ethiopia. Nat Microbiol [Internet]. 2023 Aug 28;8(10):1911–9. Available from: https://www.nature.com/articles/s41564-023-01461-4

12. Okell LC, Griffin JT, Roper C. Mapping sulphadoxine-pyrimethamine-resistant Plasmodium falciparum malaria in infected humans and in parasite populations in Africa. Sci Rep [Internet]. 2017 Dec 7;7(1):7389. Available from: 10.1038/s41598-017-06708-9

13. Matondo SI, Temba GS, Kavishe AA, Kauki JS, Kalinga A, Van Zwetselaar M, et al. High levels of sulphadoxine-pyrimethamine resistance Pfdhfr-Pfdhps quintuple mutations: a cross sectional survey of six regions in Tanzania. Malar J [Internet]. 2014 Apr 21 [cited 2025 Jul 15];13(1):152. Available from: https://pmc.ncbi.nlm.nih.gov/articles/PMC3998221/

14. Naidoo I, Roper C. Mapping ‘partially resistant’, ‘fully resistant’, and ‘super resistant’ malaria. Trends Parasitol [Internet]. 2013 Oct;29(10):505–15. Available from: 10.1016/j.pt.2013.08.002

15. Noedl H, Se Y, Schaecher K, Smith BL, Socheat D, Fukuda MM, et al. Evidence of artemisinin-resistant malaria in western Cambodia. New England Journal of Medicine [Internet]. 2008 Dec 11;359(24):2619–20. Available from: http://www.ncbi.nlm.nih.gov/pubmed/19064625

16. Dondorp AM, Nosten F, Yi P, Das D, Phyo AP, Tarning J, et al. Artemisinin Resistance in *Plasmodium falciparum* Malaria. New England Journal of Medicine [Internet]. 2009 Jul 30;361(5):455–67. Available from: http://www.ncbi.nlm.nih.gov/pubmed/25075834

17. Uwimana A, Legrand E, Stokes BH, Ndikumana JLM, Warsame M, Umulisa N, et al. Emergence and clonal expansion of *in vitro* artemisinin-resistant *Plasmodium falciparum kelch13* R561H mutant parasites in Rwanda. Nat Med [Internet]. 2020 Oct 1;26(10):1602–8. Available from: http://www.nature.com/articles/s41591-020-1005-2

18. Ndwiga L, Kimenyi KM, Wamae K, Osoti V, Akinyi M, Omedo I, et al. A review of the frequencies of Plasmodium falciparum Kelch 13 artemisinin resistance mutations in Africa. Int J Parasitol Drugs Drug Resist [Internet]. 2021 Aug;16:155–61. Available from: https://linkinghub.elsevier.com/retrieve/pii/S2211320721000282

19. Oyegbade SA, Mameh EO, Balogun DO, Aririguzoh VGO, Akinduti PA. Emerging Plasmodium falciparum K13 gene mutation to artemisinin-based combination therapies and partner drugs among malaria-infected population in sub-Saharan Africa. Parasites, Hosts and Diseases [Internet]. 2025 May 1 [cited 2025 Jul 24];63(2):109. Available from: https://pmc.ncbi.nlm.nih.gov/articles/PMC12127821/

20. Nzila AM, Mberu EK, Sulo J, Dayo H, Winstanley PA, Sibley CH, et al. Towards an Understanding of the Mechanism of Pyrimethamine-Sulfadoxine Resistance in Plasmodium falciparum: Genotyping of Dihydrofolate Reductase and Dihydropteroate Synthase of Kenyan Parasites. Antimicrob Agents Chemother [Internet]. 2000 Apr;44(4):991–6. Available from: https://journals.asm.org/doi/10.1128/AAC.44.4.991-996.2000

21. WHO. WHO Guidelines for malaria. 2024;Nov. Available from: https://app.magicapp.org/#/guideline/6812

22. Ishengoma DS, Saidi Q, Sibley CH, Roper C, Alifrangis M. Deployment and utilization of next-generation sequencing of *Plasmodium falciparum* to guide anti-malarial drug policy decisions in sub-Saharan Africa: opportunities and challenges. Malar J [Internet]. 2019 Dec 3;18(1):267. Available from: 10.1186/s12936-019-2853-4

23. Wasakul V, Verschuuren TD, Thuy-Nhien N, Booth E, Quang HH, Thang ND, et al. Genetic surveillance of *Plasmodium falciparum* populations following treatment policy revisions in the Greater Mekong Subregion. Nat Commun [Internet]. 2025 May 20;16(1):4689. Available from: https://www.nature.com/articles/s41467-025-59946-1

24. Macharia PM, Giorgi E, Noor AM, Waqo E, Kiptui R, Okiro EA, et al. Spatio-temporal analysis of Plasmodium falciparum prevalence to understand the past and chart the future of malaria control in Kenya. Malar J [Internet]. 2018 Dec 26;17(1):340. Available from: 10.1186/s12936-018-2489-9

25. Alegana VA, Macharia PM, Muchiri S, Mumo E, Oyugi E, Kamau A, et al. *Plasmodium falciparum* parasite prevalence in East Africa: Updating data for malaria stratification. Ashton R, editor. PLoS Global Public Health [Internet]. 2021 Dec 7;1(12):e0000014. Available from: https://dx.plos.org/10.1371/journal.pgph.0000014

26. National Malaria Control Programme and Ministry of Health. Kenya Malaria Strategy 2019-2023. Nairobi; 2015.

27. Fogh S, Jepsen S, Effersøe P. Chloroquine-resistant *Plasmodium falciparum* malaria in Kenya. Trans R Soc Trop Med Hyg [Internet]. 1979 Jan;73(2):228–9. Available from: https://academic.oup.com/trstmh/article-lookup/doi/10.1016/0035-9203(79)90220-7

28. Spencer HC, Masaba SC, Kiaraho D. Sensitivity of Plasmodium Falciparum Isolates to Chloroquine in Kisumu and Malindi, Kenya. Am J Trop Med Hyg [Internet]. 1982 Sep;31(5):902–6. Available from: https://www.ajtmh.org/view/journals/tpmd/31/5/article-p902.xml

29. Shretta R, Omumbo J, Rapuoda B, Snow RW. Using evidence to change antimalarial drug policy in Kenya. Tropical Medicine and International Health [Internet]. 2000 Nov;5(11):755–64. Available from: http://www.ncbi.nlm.nih.gov/pubmed/11123822

30. Amin AA, Zurovac D, Kangwana BB, Greenfield J, Otieno DN, Akhwale WS, et al. The challenges of changing national malaria drug policy to artemisinin-based combinations in Kenya. Malar J [Internet]. 2007 Dec 29;6(1):72. Available from: https://malariajournal.biomedcentral.com/articles/10.1186/1475-2875-6-72

31. EANMAT. Monitoring antimalarial drug resistance within National Malaria Control Programmes: the EANMAT experience. Tropical Medicine and International Health [Internet]. 2001 Nov;6(11):891–8. Available from: http://doi.wiley.com/10.1046/j.1365-3156.2001.00799.x

32. Rapuoda B, Ali A, Bakyaita N, Mutabingwa TK, Mwita A, Rwagacondo C, et al. The efficacy of antimalarial monotherapies, sulphadoxine-pyrimethamine and amodiaquine in East Africa: Implications for sub-regional policy. Tropical Medicine and International Health [Internet]. 2003 Oct 30;8(10):860–7. Available from: https://onlinelibrary.wiley.com/doi/10.1046/j.1360-2276.2003.01114.x

33. Halsey ES, Venkatesan M, Plucinski MM, Talundzic E, Lucchi NW, Zhou Z, et al. Capacity Development through the US President’s Malaria Initiative–Supported Antimalarial Resistance Monitoring in Africa Network. Emerg Infect Dis [Internet]. 2017 Dec;23(13):S53–6. Available from: http://www.nc.cdc.gov/eid/article/23/13/17-0366_article.htm

34. Amboko B, Stepniewska K, Macharia PM, Machini B, Bejon P, Snow RW, et al. Trends in health workers’ compliance with outpatient malaria case-management guidelines across malaria epidemiological zones in Kenya, 2010–2016. Malar J [Internet]. 2020 Dec 11;19(1):406. Available from: 10.1186/s12936-020-03479-z

35. Tricco AC, Lillie E, Zarin W, O’Brien KK, Colquhoun H, Levac D, et al. PRISMA Extension for Scoping Reviews (PRISMA-ScR): Checklist and Explanation. Ann Intern Med [Internet]. 2018 Oct 2;169(7):467–73. Available from: https://www.acpjournals.org/doi/10.7326/M18-0850

36. Ouzzani M, Hammady H, Fedorowicz Z, Elmagarmid A. Rayyan—a web and mobile app for systematic reviews. Syst Rev [Internet]. 2016 Dec 5;5(1):210. Available from: http://systematicreviewsjournal.biomedcentral.com/articles/10.1186/s13643-016-0384-4

37. Aurrecoechea C, Brestelli J, Brunk BP, Dommer J, Fischer S, Gajria B, et al. PlasmoDB: a functional genomic database for malaria parasites. Nucleic Acids Res. 2009 Jan;37(Database issue):D539-43.

38. Bahl A, Brunk B, Crabtree J, Fraunholz MJ, Gajria B, Grant GR, et al. PlasmoDB: the Plasmodium genome resource. A database integrating experimental and computational data. Nucleic Acids Res. 2003 Jan 1;31(1):212–5.

39. Fidock DA, Nomura T, Talley AK, Cooper RA, Dzekunov SM, Ferdig MT, et al. Mutations in the P. falciparum digestive vacuole transmembrane protein PfCRT and evidence for their role in chloroquine resistance. Mol Cell. 2000;6(4):861–71.

40. Djimdé A, Doumbo OK, Cortese JF, Kayentao K, Doumbo S, Diourté Y, et al. A Molecular Marker for Chloroquine-Resistant Falciparum Malaria. New England Journal of Medicine [Internet]. 2001 Jan 25;344(4):257–63. Available from: http://www.nejm.org/doi/abs/10.1056/NEJM200101253440403

41. Sidhu ABS, Verdier-Pinard D, Fidock DA. Chloroquine Resistance in Plasmodium falciparum Malaria Parasites Conferred by pfcrt Mutations. Science (1979) [Internet]. 2002 Oct 4;298(5591):210–3. Available from: http://www.sciencemag.org/content/298/5591/210.short

42. Foote SJ, Kyle DE, Martin RK, Oduola AMJ, Forsyth K, Kemp DJ, et al. Several alleles of the multidrug-resistance gene are closely linked to chloroquine resistance in *Plasmodium falciparum*. Nature [Internet]. 1990 May;345(6272):255–8. Available from: http://www.nature.com/articles/345255a0

43. Peterson DS, Walliker D, Wellems TE. Evidence that a point mutation in dihydrofolate reductase-thymidylate synthase confers resistance to pyrimethamine in *falciparum* malaria. Proceedings of the National Academy of Sciences [Internet]. 1988 Dec 1;85(23):9114–8. Available from: http://www.pnas.org/cgi/doi/10.1073/pnas.85.23.9114

44. Foote SJ, Galatis D, Cowman AF. Amino acids in the dihydrofolate reductase-thymidylate synthase gene of *Plasmodium falciparum* involved in cycloguanil resistance differ from those involved in pyrimethamine resistance. Proceedings of the National Academy of Sciences [Internet]. 1990 Apr 1;87(8):3014–7. Available from: http://www.pnas.org/cgi/doi/10.1073/pnas.87.8.3014

45. Basco LK, de Pécoulas PE, Wilson CM, Le Bras J, Mazabraud A. Point mutations in the dihydrofolate reductase-thymidylate synthase gene and pyrimethamine and cycloguanil resistance in *Plasmodium falciparum*. Mol Biochem Parasitol [Internet]. 1995 Jan;69(1):135–8. Available from: https://linkinghub.elsevier.com/retrieve/pii/0166685194002074

46. Triglia T, Wang P, Sims PFG, Hyde JE, Cowman AF. Allelic exchange at the endogenous genomic locus in Plasmodium falciparum proves the role of dihydropteroate synthase in sulfadoxine-resistant malaria. EMBO J [Internet]. 1998 Jul 15;17(14):3807–15. Available from: http://emboj.embopress.org/cgi/doi/10.1093/emboj/17.14.3807

47. WHO. Artemisinin resistance and artemisinin-based combination therapy efficacy. World Health Organisation. https://www.who.int/malaria/publications/atoz/artemisinin-resistance-august2018/en/; 2018.

48. Witkowski B, Duru V, Khim N, Ross LS, Saintpierre B, Beghain J, et al. A surrogate marker of piperaquine-resistant *Plasmodium falciparum* malaria: a phenotype–genotype association study. Lancet Infect Dis [Internet]. 2017 Feb;17(2):174–83. Available from: http://linkinghub.elsevier.com/retrieve/pii/S1473309916304157

49. Bopp S, Magistrado P, Wong W, Schaffner SF, Mukherjee A, Lim P, et al. Plasmepsin II–III copy number accounts for bimodal piperaquine resistance among Cambodian *Plasmodium falciparum*. Nat Commun [Internet]. 2018 Dec 2;9(1):1769. Available from: 10.1038/s41467-018-04104-z

50. Demas AR, Sharma AI, Wong W, Early AM, Redmond S, Bopp S, et al. Mutations in *Plasmodium falciparum* actin-binding protein coronin confer reduced artemisinin susceptibility. Proceedings of the National Academy of Sciences [Internet]. 2018 Dec 11;115(50):12799–804. Available from: http://www.pnas.org/lookup/doi/10.1073/pnas.1812317115

51. Dahlström S, Ferreira PE, Veiga MI, Sedighi N, Wiklund L, Mårtensson A, et al. *Plasmodium falciparum* Multidrug Resistance Protein 1 and Artemisinin-Based Combination Therapy in Africa. J Infect Dis [Internet]. 2009 Nov;200(9):1456–64. Available from: https://academic.oup.com/jid/article-lookup/doi/10.1086/606009

52. Henriques G, Hallett RL, Beshir KB, Gadalla NB, Johnson RE, Burrow R, et al. Directional Selection at the *pfmdr1, pfcrt, pfubp1*, and *pfap2mu* Loci of *Plasmodium falciparum* in Kenyan Children Treated With ACT. J Infect Dis [Internet]. 2014 Dec 15;210(12):2001–8. Available from: http://jid.oxfordjournals.org/lookup/doi/10.1093/infdis/jiu358

53. Henrici RC, Sutherland CJ. Alternative pathway to reduced artemisinin susceptibility in *Plasmodium falciparum*. Proceedings of the National Academy of Sciences [Internet]. 2018;201818287. Available from: http://www.pnas.org/content/early/2018/11/27/1818287115

54. Amato R, Lim P, Miotto O, Amaratunga C, Dek D, Pearson RD, et al. Genetic markers associated with dihydroartemisinin–piperaquine failure in *Plasmodium falciparum* malaria in Cambodia: a genotype–phenotype association study. Lancet Infect Dis [Internet]. 2017 Feb;17(2):164–73. Available from: http://linkinghub.elsevier.com/retrieve/pii/S1473309916304091

55. Thiers BH. Resistance of *Plasmodium falciparum* Field Isolates to In-Vitro Artemether and Point Mutations of the SERCA-Type PfATPase6. Yearbook of Dermatology and Dermatologic Surgery [Internet]. 2006 Jan;2006:155–6. Available from: https://linkinghub.elsevier.com/retrieve/pii/S0093361908701102

56. Pillai DR, Lau R, Khairnar K, Lepore R, Via A, Staines HM, et al. Artemether resistance in vitro is linked to mutations in PfATP6 that also interact with mutations in PfMDR1 in travellers returning with *Plasmodium falciparum* infections. Malar J [Internet]. 2012 Dec 27;11(1):131. Available from: https://malariajournal.biomedcentral.com/articles/10.1186/1475-2875-11-131

57. Ariey F, Witkowski B, Amaratunga C, Beghain J, Langlois AC, Khim N, et al. A molecular marker of artemisinin-resistant *Plasmodium falciparum* malaria. Nature [Internet]. 2014 Jan 18;505(7481):50–5. Available from: http://www.nature.com/doifinder/10.1038/nature12876

58. Amambua-Ngwa A, Jeffries D, Amato R, Worwui A, Karim M, Ceesay S, et al. Consistent signatures of selection from genomic analysis of pairs of temporal and spatial *Plasmodium falciparum* populations from The Gambia. Sci Rep [Internet]. 2018 Dec 26;8(1):9687. Available from: http://www.nature.com/articles/s41598-018-28017-5

59. Miotto O, Amato R, Ashley EA, MacInnis B, Almagro-Garcia J, Amaratunga C, et al. Genetic architecture of artemisinin-resistant *Plasmodium falciparum*. Nat Genet [Internet]. 2015 Mar 19;47(3):226–34. Available from: http://www.nature.com/doifinder/10.1038/ng.3189

60. Haddaway NR, Page MJ, Pritchard CC, McGuinness LA. PRISMA2020: An R package and Shiny app for producing PRISMA 2020-compliant flow diagrams, with interactivity for optimised digital transparency and Open Synthesis. Campbell Systematic Reviews [Internet]. 2022 Jun 27;18(2). Available from: https://onlinelibrary.wiley.com/doi/10.1002/cl2.1230

61. Osoti V, Wamae K, Ndwiga L, Gichuki PM, Okoyo C, Kepha S, et al. Detection of low frequency artemisinin resistance mutations, C469Y, P553L and A675V, and fixed antifolate resistance mutations in asymptomatic primary school children in Kenya. BMC Infect Dis [Internet]. 2025 Jan 16;25(1):73. Available from: https://bmcinfectdis.biomedcentral.com/articles/10.1186/s12879-025-10462-z

62. Taylor SM, Parobek CM, DeConti DK, Kayentao K, Coulibaly SO, Greenwood BM, et al. Absence of Putative Artemisinin Resistance Mutations Among *Plasmodium falciparum* in Sub-Saharan Africa: A Molecular Epidemiologic Study. J Infect Dis [Internet]. 2015 Mar 1;211(5):680–8. Available from: https://academic.oup.com/jid/article/211/5/680/2918011

63. Jeang B, Zhong D, Lee MC, Atieli H, Yewhalaw D, Yan G. Molecular surveillance of Kelch 13 polymorphisms in *Plasmodium falciparum* isolates from Kenya and Ethiopia. Malar J [Internet]. 2024 Jan 29;23(1):36. Available from: 10.1186/s12936-023-04812-y

64. Maniga JN, Samuel M, John O, Rael M, Muchiri JN, Bwogo P, et al. Novel *Plasmodium falciparum k13* gene polymorphisms from Kisii County, Kenya during an era of artemisinin-based combination therapy deployment. Malar J [Internet]. 2023 Mar 9;22(1):87. Available from: 10.1186/s12936-023-04517-2

65. Makau M, Kanoi BN, Mgawe C, Maina M, Bitshi M, Too EK, et al. Presence of *Plasmodium falciparum* strains with artemisinin-resistant *k13* mutation C469Y in Busia County, Western Kenya. Trop Med Health [Internet]. 2024 Oct 18;52(1):72. Available from: http://www.ncbi.nlm.nih.gov/pubmed/39425178 %0A http://www.pubmedcentral.nih.gov/articlerender.fcgi?artid=PMC11488207

66. Omedo I, Bartilol B, Kimani D, Gonçalves S, Drury E, Rono MK, et al. Spatio-temporal distribution of antimalarial drug resistant gene mutations in a Plasmodium falciparum parasite population from Kilifi, Kenya: A 25-year retrospective study. Wellcome Open Res [Internet]. 2022 Feb 8;7:45. Available from: https://wellcomeopenresearch.org/articles/7-45/v1

67. van Eijk AM, Larsen DA, Kayentao K, Koshy G, Slaughter DEC, Roper C, et al. Effect of Plasmodium falciparum sulfadoxine-pyrimethamine resistance on the effectiveness of intermittent preventive therapy for malaria in pregnancy in Africa: a systematic review and meta-analysis. Lancet Infect Dis [Internet]. 2019 Mar;3099(18):1–11. Available from: 10.1016/S1473-3099(18)30732-1

68. Sanchez CP, McLean JE, Rohrbach P, Fidock DA, Stein WD, Lanzer M. Evidence for a *Pf*crt-associated chloroquine efflux system in the human malarial parasite *Plasmodium falciparum*. Biochemistry [Internet]. 2005 Jul;44(29):9862–70. Available from: https://pubs.acs.org/doi/10.1021/bi050061f

69. Duraisingh MT, Cowman AF. Contribution of the pfmdr1 gene to antimalarial drug-resistance. Acta Trop. 2005;94(3 SPEC. ISS.):181–90.

70. Duraisingh MT, Roper C, Walliker D, Warhurst DC. Increased sensitivity to the antimalarials mefloquine and artemisinin is conferred by mutations in the pfmdr1 gene of Plasmodium falciparum. Mol Microbiol [Internet]. 2000 May;36(4):955–61. Available from: http://doi.wiley.com/10.1046/j.1365-2958.2000.01914.x

71. Reed MB, Saliba KJ, Caruana SR, Kirk K, Cowman AF. *Pgh1* modulates sensitivity and resistance to multiple antimalarials in *Plasmodium falciparum*. Nature [Internet]. 2000 Feb 24;403(6772):906–9. Available from: http://www.nature.com/doifinder/10.1038/35002615

72. Mwai L, Kiara SM, Abdirahman A, Pole L, Rippert A, Diriye A, et al. In Vitro Activities of Piperaquine, Lumefantrine, and Dihydroartemisinin in Kenyan Plasmodium falciparum Isolates and Polymorphisms in Pfcrt and Pfmdr1. Antimicrob Agents Chemother [Internet]. 2009 Dec 1 [cited 2014 Dec 1];53(12):5069–73. Available from: http://www.pubmedcentral.nih.gov/articlerender.fcgi?artid=2786317&tool=pmcentrez&rendertype=abstract

73. Pickard AL, Wongsrichanalai C, Purfield A, Kamwendo D, Emery K, Zalewski C, et al. Resistance to Antimalarials in Southeast Asia and Genetic Polymorphisms in pfmdr1. Antimicrob Agents Chemother [Internet]. 2003 Aug;47(8):2418–23. Available from: https://journals.asm.org/doi/10.1128/AAC.47.8.2418-2423.2003

74. Sidhu ABS, Valderramos SG, Fidock D a. pfmdr1 mutations contribute to quinine resistance and enhance mefloquine and artemisinin sensitivity in Plasmodium falciparum. Mol Microbiol. 2005;57:913–26.

75. Sidhu ABS, Uhlemann AC, Valderramos SG, Valderramos JC, Krishna S, Fidock D a. Decreasing pfmdr1 copy number in plasmodium falciparum malaria heightens susceptibility to mefloquine, lumefantrine, halofantrine, quinine, and artemisinin. J Infect Dis. 2006;194:528–35.

76. Tessema SK, Inzaule SC, Christoffels A, Kebede Y, de Oliveira T, Ouma AEO, et al. Accelerating genomics-based surveillance for COVID-19 response in Africa. Lancet Microbe [Internet]. 2020 Oct;1(6):e227–8. Available from: https://linkinghub.elsevier.com/retrieve/pii/S2666524720301178

